# FARFOOD: A database of potential interactions between food compounds and drugs

**DOI:** 10.1101/2024.11.22.24317802

**Authors:** Ana María Nevado-Bulnes, Dixan Agustin Benitez, Guadalupe Cumplido-Laso, Jose María Carvajal-González, Sonia Mulero-Navarro, Angel-Carlos Román

**Affiliations:** Servicio Extremeño de Salud (SES), C/Charcón s/n, 10350 – Almaraz (Spain); Departamento de Bioquímica y Biología Molecular y Genética, Universidad de Extremadura. Avda de Elvas s/n, 06071 – Badajoz (Spain)

**Keywords:** Food-Drug Interactions, in silico modeling, lisinopril, bupropion, allopurinol

## Abstract

Nutrition is a fundamental aspect in human life development, both from a socio-economic and medical perspective. In recent years, new personalized approaches have been added to the biochemical study of nutrients. These approaches consider both the effect of foods on the body and the role of genes in metabolizing or digesting different nutrients. Although drug-food interactions have been known for decades, there is a lack of studies that address these processes in a comprehensive way, using structural and computational biochemistry techniques. In this paper we develop a method to predict potential interactions between foods and drugs based on the structural similarity between food compounds and medications. Our results have produced a database and an app to consult potential interactions between drugs and foods that we have called FARFOOD. Additionally, we validated two of these potential interactions with widely used drugs (lisinopril and bupropion) through structural docking between the ligand protein and the food compounds that are structurally similar to the drug. Moreover, patient surveys are used in the lisinopril and bupropion cases in addition to allopurinol to assess the possible effects of the potentially interacting foods on the symptoms of the conditions for which the medication is prescribed. In summary, this manuscript presents an interesting computational resource for predictive food-drug interaction analysis.

## Introduction

Food-drug interactions are a crucial yet often underappreciated aspect of patient care and therapeutic efficacy. The ingestion of certain foods can significantly alter the pharmacokinetics and pharmacodynamics of drugs, impacting their absorption, metabolism, and overall effectiveness (Genser, 2008; Z. Hu et al., 2005; Won et al., 2012). These interactions may lead to reduced therapeutic effects or, conversely, to toxicity, making it essential for healthcare providers and patients to understand the role diet plays in the success or failure of drug regimens (Chen et al., 2012; Koziolek et al., 2019; Won et al., 2012). With the rise of personalized medicine, understanding food-drug interactions is more important than ever, as individual genetic variations can influence how drugs and food components interact within the body (Roy et al., 2022; Wei et al., 2024). Despite the well-documented impact of food-drug interactions on clinical outcomes, systematic research and data collection in this area are limited. Most current guidelines focus on well-known interactions (Bailey & Dresser, 2004; Hukkinen et al., 1995; Mertens-Talcott et al., 2006; Seden et al., 2012; Sica, 2006), but there are many unexplored interactions that could affect patient safety and drug efficacy, especially in populations consuming diverse diets.

This complexity of food-drug interactions is attributed to various factors, including the biochemical properties of both drugs and food compounds. For instance, foods containing high levels of specific nutrients, such as calcium or certain fibers, can inhibit the absorption of certain medications (Aznar-Lou et al., 2019; Deng et al., 2017; Neuvonen, 1976; Stielow et al., 2023), while others, like those containing grapefruit compounds, can potentiate drug effects by inhibiting cytochrome P450 enzymes (Mertens-Talcott et al., 2006; Seden et al., 2012). A potential manner for detecting food-drug interactions is based on structural similarity between food compounds and drugs, as it has been done with drug-drug interactions (Takeda et al., 2017; Vilar et al., 2012). Structural similarity implies that certain bioactive compounds in foods may share similar chemical structures with drug molecules, potentially leading them to bind to the same target sites in the body. This structural resemblance can result in competition for binding sites, altering the pharmacokinetics or pharmacodynamics of drugs. Understanding these structural relationships is increasingly important for creating dietary guidelines tailored to individual pharmacological needs, especially as dietary diversity and self-medication continue to rise globally.

Structural similarity-based predictions provide a unique perspective for analyzing interactions that might not be evident through traditional pharmacokinetic studies alone. Computational approaches such as molecular docking (Agu et al., 2023), quantitative structure-activity relationship (QSAR) modeling (Zaki et al., 2021), and machine learning (Roy et al., 2022) can be applied to screen food compounds against drug targets. For example, certain polyphenols in fruits and vegetables have structural motifs resembling common pharmacophores in drug molecules, allowing them to interact with cytochrome P450 enzymes, potentially inhibiting or inducing drug metabolism (Kimura et al., 2010). Such findings remark the need for systematic screening of food compounds using computational models, which could help predict and ultimately prevent adverse interactions. This highlights the potential of using structural similarity as a predictive tool in food-drug interaction research, where identifying molecular similarities between food and drug compounds can lead to proactive dietary or medicament doses recommendations.

The main aim of this manuscript is to develop a comprehensive computational resource for potential interactions between drugs and food compounds based on structural similarity. Other recent approaches have used gene expression and other sources for the development of similar databases (Lacruz-Pleguezuelos et al., 2023). We considered as hypothesis that if two molecules have a very similar structure, then they might interact with the same protein targets. We validated some of these interactions using (i) *in silico* docking experiments that showed how both the drug and the food compound can bind to the same protein region and (ii) patient surveys in which we correlate disease parameters with the consumption of foods that contain the potentially interacting compound.

## Materials and Methods

### Input food compound and drug data

For food-related data, we utilized version 1.0 of FooDB (www.foodb.ca) , a comprehensive, publicly funded database supported by the Canadian Institutes of Health. FooDB provides extensive information on the biology and chemistry of food components, including macronutrients, micronutrients, and attributes like color, aroma, flavor, and texture. Each food component entry includes over 100 fields covering nomenclature, structural and physicochemical properties, food sources, concentrations, physiological effects, and potential health impacts (Sanchez-Ruiz & Colmenarejo, 2021). For our structural similarity analysis, we used only the structural data (compound ID and SMILES format), while food source and concentration data were applied in later analyses and for the app development. For drug information, we employed version 15 of CHEMBL (www.ebi.ac.uk/chembl), a manually curated database of bioactive molecules maintained by the European Bioinformatics Institute (Gaulton et al., 2011). CHEMBL includes structural data as well as physicochemical and pharmacological properties for each molecule, such as molecular weight, Lipinski parameters, ADME properties, and molecular targets. We filtered CHEMBL entries to include only molecules approved or in late-stage studies, narrowing the dataset from over two million entries to approximately 10,000 compounds. For subsequent analyses and the app, we included each drug’s generic name and physicochemical and pharmacological characteristics.

### Structural comparison between food compounds and drugs

To identify potential interactions between food compounds and drugs, we conducted a structural similarity analysis between each food compound and drug. This one-to-one comparison across over 70,000 food compounds and more than 10,000 drugs results in over 700 million possible pairwise comparisons. Due to the scale, these comparisons were automated using a computational algorithm to process, compare, and analyze each compound-drug pair. To facilitate this process, we developed a PERL script that utilizes an external software tool, Open Babel (www.openbabel.org) (O’Boyle et al., 2011), to generate a list of food compound/drug pairs with structural similarity above a defined threshold. For similarity measurement, we employed the Tanimoto index (Tanimoto, 1958), a widely-used metric in cheminformatics based on molecular fingerprints—a binary encoding of the molecular structure (Willett et al., 1986). Each fingerprint represents the presence or absence of specific substructures within a molecule using binary bits. By setting a threshold, we identified molecule pairs that met or exceeded this similarity score, with higher threshold values indicating greater structural similarity.

### Design of FARFOOD database and app

The molecule pairs with Tanimoto index > 0.7 were selected, and the information obtained from the FooDB and CHEMBL databases was stored as data tables in MATLAB. To generalize and simplify access to the FARFOOD database, we developed a cross-platform app using MATLAB’s Application Designer. The app is compatible with Windows, Linux, and macOS, though it requires users to install the free MATLAB Runtime environment. The app is distributed as an executable file that can be downloaded from http://www.github.com/acroman. The FARFOOD app provides access to the database information, allowing users to explore drug-food compound interactions, as well as detailed data from CHEMBL and FooDB. The app’s interface prioritizes simplicity, featuring a search bar at the top, a dropdown menu to select either "CHEMBL ID" or "CHEMBL name," and a search button for easy querying. The app is centered around drugs, enabling users to quickly identify potential food interactions for any drug of interest. To enrich the interaction data, the app links interaction results with additional details on compounds, including the foods containing each compound and their concentrations. This information is shown in a lower app window when the user clicks on a compound, enabling a comprehensive view of food sources and concentrations for each compound involved in a drug interaction.

### Molecular docking

To validate potential interactions between structurally similar food compounds and drugs, we employed a molecular docking approach (Shoichet, 2004). Docking analyses require the structure of the target protein and the potential ligand as input, producing one or more bound conformations of the ligand-protein complex as output. We selected the target protein for each drug as identified in the FARFOOD database, which draws on CHEMBL for target information. PDB files for each protein were downloaded from the Protein Data Bank (Berman et al., 2000). For ligands, we used both the drug (as a positive control) and the structurally similar food compound. Drug structures were obtained from CHEMBL, while food compound structures were retrieved from FooDB and converted from SMILES to PDB format using Open Babel. Docking was performed using DockThor (https://www.dockthor.lncc.br/v2/) (Guedes et al., 2024), a public server supporting interactive docking. Default conditions, including blind docking, were applied to ensure comprehensive exploration of binding sites.

### Patient surveys

Patients with specific drug prescription from the regional healthcare area of Almaraz were contacted by phone. They were asked for their habits of food consumption (using foods that contain potentially interacting compounds as well as other –control-types of food). These results were normalized using the following scale: 0-never; 1-sporadically, once in months; 2-at least once per week; 3-daily consumption. In addition, they were asked for specific symptoms or characteristics about the diseases related to the prescription. A minimum of 20 patients were asked for each drug analyzed, and the demographic data (age and sex) can be consulted in Table 1. The study protocol was reviewed by the Ethics Board of the University of Extremadura, and approved with the reference number 167/2023.

**Table 1.** Demographic data of patients used in the study.

| **Drug** | **Sex (Female %)** | **Age (Mean/SD years)** |
| --- | --- | --- |
| Lisinopril | 48% | 75.2/12.3 |
| Bupropion | 55% | 51.1/20.4 |
| Allopurinol | 22% | 73.1/18.8 |

## Results

### Design of a database of potential interactions between drug and food compounds using structural similarity

First we designed a workflow that detected potential interactions between drug and food compounds based on the structural similarity of each molecule (Figure 1A). Specifically, we compared the structure of each drug present in CHEMBL database (N=10,650) with each food compound present in FooDB database (N=70,477). We retrieved the pairwise relations that presented a Tanimoto Index equal or above 0.7, resulting in 657,219 potential interactions between drugs and food compounds. We compiled specific information from both databases in a MATLAB application that can be freely used (Figure 1B). With this application, a user can search for a specific drug, and the database will return the specific compounds with structural similarity (Tanimoto Index equal or above 0.7), the food in which this compound has been detected and its relative concentration, if known. As expected, the number of drugs that present potential interactions with food compounds decreases if we increase the Tanimoto Index threshold (Figure 1C). More than 2,500 drugs in the CHEMBL database present structural similarities with a Tanimoto Index threshold of 0.7, while more than 1,000 drugs still present structural similarities with a threshold of 0.9. The specificity of the potential interactions that were predicted is supported because most of the drugs presented only 1-5 interactions with compounds (Figure 1D). This data showed that a relevant number of known drugs presented structural similarity to food compounds and their specificity suggested that they might become candidates for a source of food-drug interactions.

**Figure 1.**
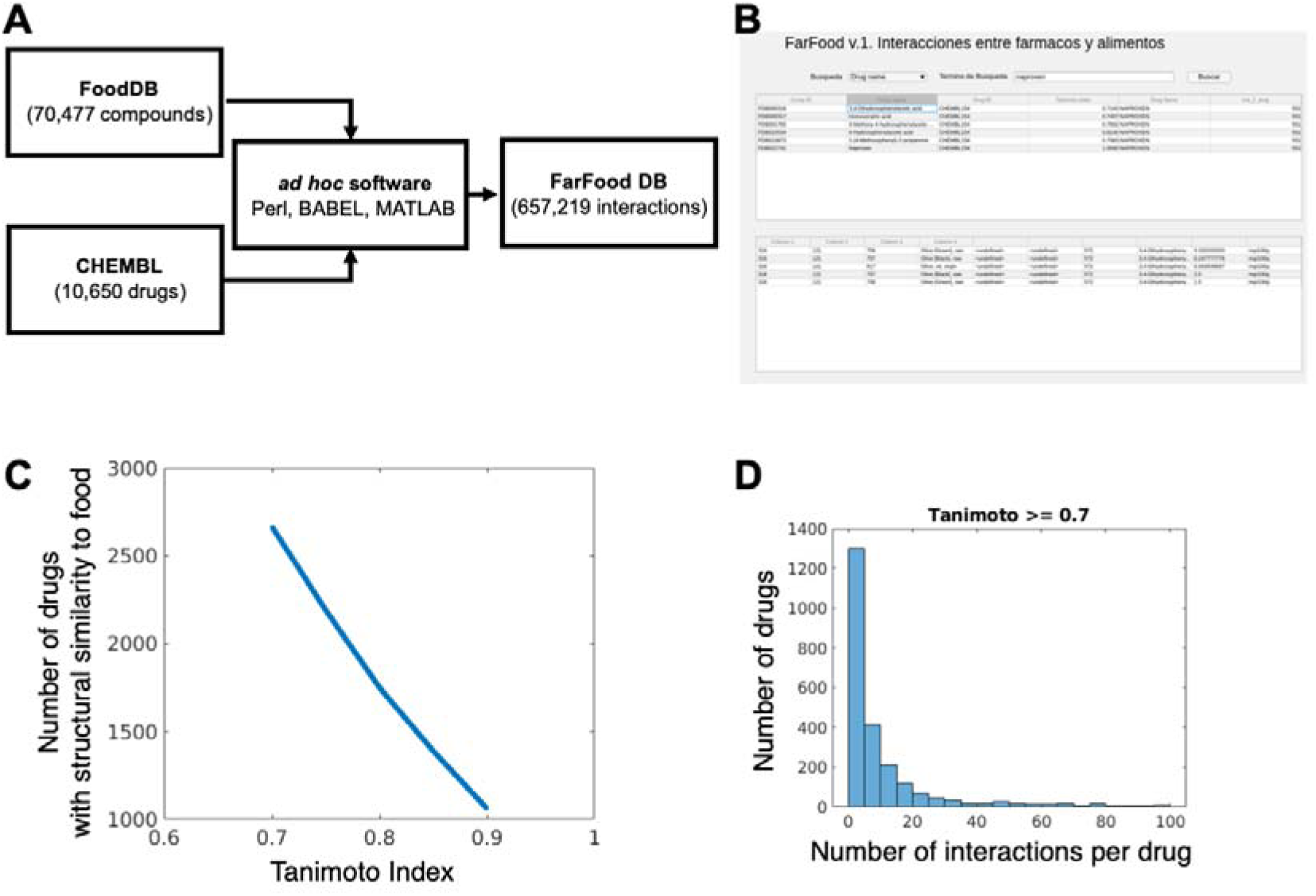
**(A)** Scheme of the food-wide and drug-wide analysis to detect structural similarities between drugs and food compounds. **(B)** Snapshot of the FARFOOD application available at https://github.com/angelcroman. **(C)** Number of drugs with potential food interactions depending on the Tanimoto Index. **(D)** Histogram representing the different number of food compounds that present structural similarity to a specific drug.

### Study of the pharmacological and chemical properties of candidate drugs

We explored further the drugs that presented structural similarity with food compounds, using some of their pharmacological and chemical properties that were contained in CHEMBL database. The majority (56%) of these drugs were synthetic small molecules, while 35% of them were from natural origin, a property that could be linked to their similarity to natural food compounds (Figure 2A). The structure of these drugs mainly presented single steroisomerism (47%), with a representative percentage (27%) being achiral molecules (Figure 2B). As this study focused in the detection of potential interactions between drugs and food compounds, it was very relevant for us to assess the percentage of these drugs that are orally administered, as they might directly interact with the food compounds. We observed that 12% of the drugs that presented structural similarity with food compounds were orally administered (Figure 2C). In addition, one of the main classic methods to determine the potential oral bioavailability of a drug is the Rule of Lipinski or Rule of 5, that characterize the pharmacokinetic properties of a drug. In this case, 62% of the drugs with potential food interactions fulfilled this Rule (Figure 2D). Finally, it was also interesting to assess the novelty of these drugs, but a low percentage (1%) of them were the first-of-class drug on its area (Figure 2E). This exploratory study confirms that several drug candidates can physically share the space with food compounds, leading to a more severe interaction in the digestive system.

**Figure 2.**
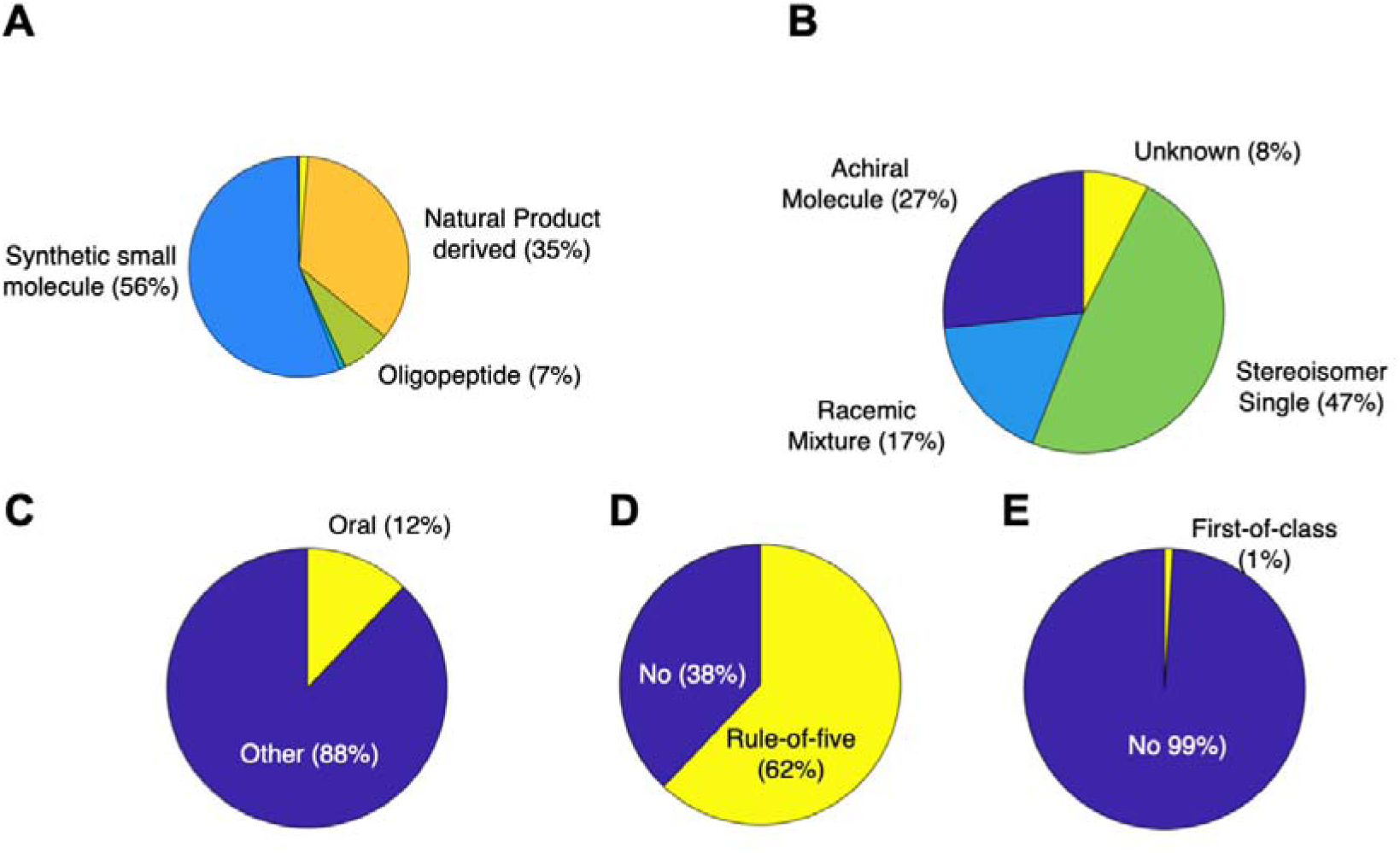
Pie charts categorizing the drugs that presented structural similarities with food compounds depending on **(A)** the molecular origin of the drug, **(B)** the structural properties of the drug, **(C)** the administration (oral or not) of the drug, **(D)** the fulfillment of the Rule-of-five or Rule of Lipinski, and **(E)** the novelty of the drug among its class.

### Study case of lisinopril and its potential interaction with cyclosquamosin A

Lisinopril is a peptidyl dipeptidase inhibitor classified under ACE inhibitors (angiotensin-converting enzyme inhibitors) (Chase & Sutton, 1989; Lancaster & Todd, 1988). It functions by competitively inhibiting the ACE enzyme, which catalyzes the conversion of angiotensin I to the vasoconstrictor peptide angiotensin II. Angiotensin II also stimulates aldosterone secretion by the adrenal cortex. Inhibiting ACE reduces angiotensin II levels, leading to decreased vasopressor activity and reduced aldosterone secretion, which may increase serum potassium levels. Through this mechanism, lisinopril acts as a potent vasodilator for both veins and arteries, producing prolonged hypotensive effects. Lisinopril is primarily indicated for hypertension, offering advantages over other antihypertensives as it does not interfere with carbohydrate, lipid, or uric acid metabolism. Additionally, it is used to treat conditions such as heart failure, acute myocardial infarction, and diabetic nephropathy. A common side effect is a persistent dry cough, which may lead some patients to discontinue treatment (Y. Hu et al., 2023). According to DrugStats, lisinopril was the fourth most prescribed drug in the United States in 2020, with 88,597,017 prescriptions for 19,816,361 patients.

In terms of potential food-drug interactions, the FARFOOD database estimated its structural similarity with cyclosquamosin A (Figure 3A, top). Cyclosquamosin A is a compound found in tropical fruits such as cherimoya (Yang et al., 2008). Interestingly, other compounds from its same family showed vasorelaxant and anti-inflammatory activities (Morita et al., 2006; Yang et al., 2008). Since the molecular target of lisinopril (ACE) is known, we performed docking analyses to examine whether cyclosquamosin A binds to the same site. It is shown that both compounds have affinity for the same binding region on ACE (Figure 3A, middle). A closer look at the binding pocket (Figure 3A, bottom) reveals that both compounds occupy the same pocket on ACE, with cyclosquamosin A forming even more interactions. This supports the possibility of an interaction between the two molecules. In order to confirm this, we studied if the consumption of tropical fruits (with content of cyclosquamosin A) in lisinopril patients was associated to the presence of past myocardial infarctions (Figure 3B, top). We statistically observed that the consumption of tropical fruits was associated to the absence of myocardial infarctions, while this association was not observed in the case of other fruit consumption (Figure 3B, bottom). This result supports the idea of a potential interaction between cyclosquamosin A and lisinopril.

**Figure 3.**
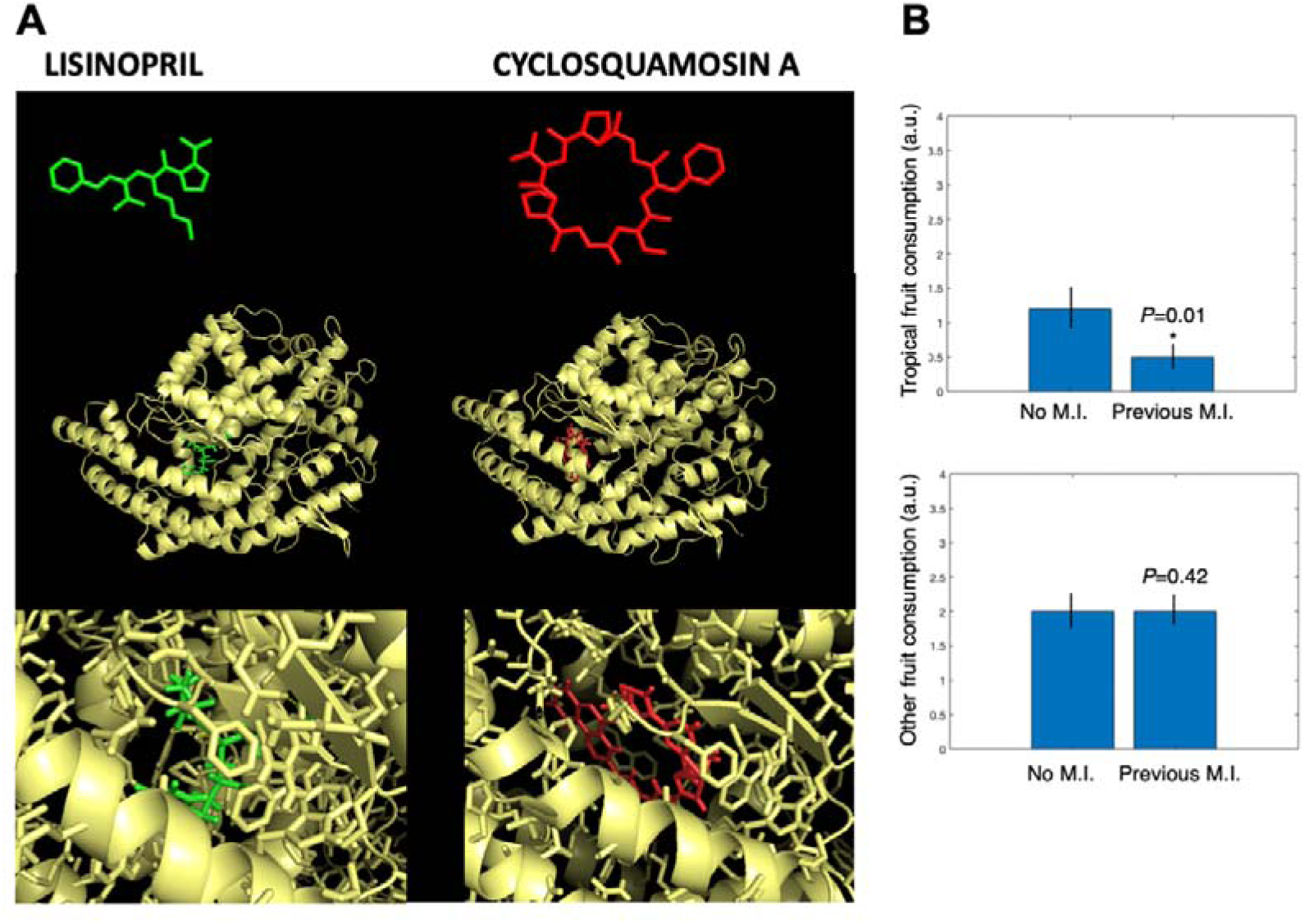
**(A)** Structures of lisinopril and cyclosquamosin A (top), as well as the docking results of both compounds against ACE (middle, bottom). **(B)** Results from patient surveys related to tropical (top) or other (bottom) fruit consumption and the presence of myocardial infarctions (M.I).

### Study case of bupropion and its potential interaction with hydroxybupropion

Bupropion is classified under the therapeutic category of other antidepressants and is primarily indicated for major depressive episodes (Dhillon et al., 2008; Patel et al., 2016). Its mechanism of action involves the selective inhibition of catecholamine reuptake, specifically norepinephrine and dopamine, with minimal effect on serotonin reuptake and no inhibition of monoamine oxidase (Stahl et al., 2004). This selective inhibition leads to an increase in neurotransmitters within the synaptic cleft, which is associated with improved mood and a sense of well-being. Bupropion is also approved as a smoking cessation aid for patients with nicotine dependence, although the exact mechanism for this effect is not fully understood . It is believed to involve noradrenergic and/or dopaminergic pathways (Simon et al., 2004). According to DrugStats, bupropion was the 18th most prescribed medication in the United States in 2020, with 28,889,368 prescriptions for 5,801,282 patients.

Structural similarity analysis revealed that only hydroxybupropion, a metabolite of bupropion (Connarn et al., 2016), shares a significant structural resemblance with bupropion itself (Tanimoto index = 0.875; Figure 4A, top). Following FooDB, hydroxybupropion has been detected, although not quantified, in various foods such as black gram beans (*Vigna mungo*), chicory leaves (*Cichorium intybus var. foliosum*), red currants *(Ribes rubrum*), and European chestnuts (*Castanea sativa*). Given that the molecular target of bupropion is known (the dopamine transporter, DAT) (Nemeroff & Owens, 2009), a docking analysis was conducted to assess whether hydroxybupropion binds to the same region on DAT using the DockThor server. As shown in figure 4A (middle), both compounds bound to the same region of DAT, supporting the hypothesis that hydroxybupropion may interfere with the effect of bupropion if consumed through foods containing this metabolite. A closer examination of the binding site (Figure 4A, bottom) illustrates the significant overlap in the binding positions of both molecules, further supporting the potential for interaction. Previous works suggested that hydroxybupropion can also reduce depression (Damaj et al., 2004), so we studied if the patterns in hydroxybupropion-containing food consumption can interact with parameters related to bupropion prescription. In this case, we observed that the consumption of chestnuts (food that contains hydroxybupropion) is statistically associated to mild instead of severe depression in the bupropion patients (Figure 4B, top). In the case of a control food that does not contain hydroxybupropion, as milk, we did not observe the same trend (Figure 4B, bottom). As in with lisinopril, this survey data points to an interaction between the drug and the food compound, or in this case bupropion and chestnut consumption.

**Figure 4.**
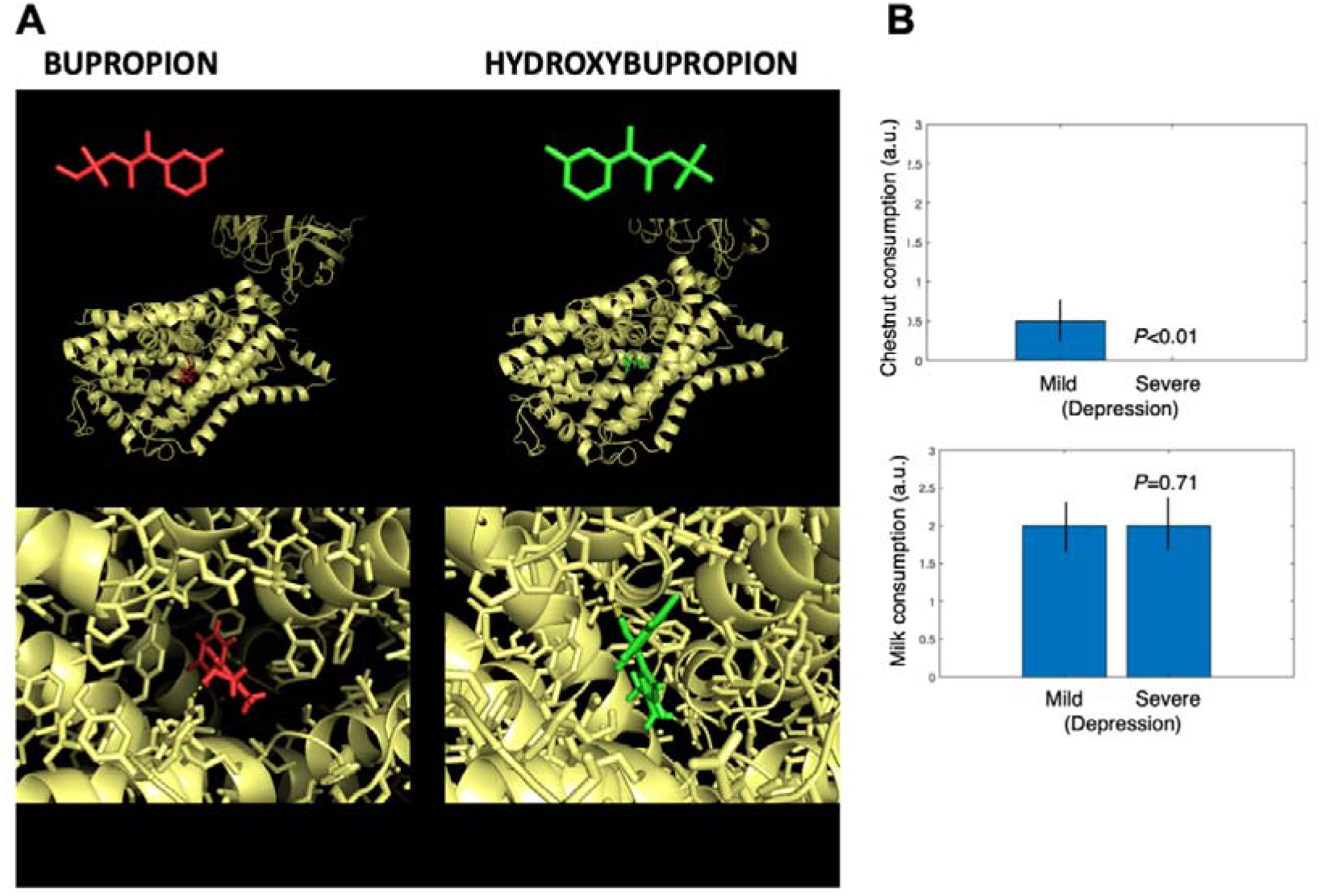
**(A)** Structures of bupropion and hydroxybupropion (top), as well as the docking results of both compounds against DAT (middle, bottom). **(B)** Results from patient surveys related to chestnut (top) or milk (bottom) consumption and the presence of episodes of mild or severe depression.

### Study case of allopurinol and its potential interaction with oxypurinol

Allopurinol is classified as a uric acid production inhibitor, working primarily by inhibiting xanthine oxidase (Gibson et al., 1982; Seth et al., 2014). This enzyme is responsible for converting hypoxanthine to xanthine and xanthine to uric acid. By blocking this pathway, allopurinol reduces uric acid levels both in plasma and urine (Gibson et al., 1982; Seth et al., 2014). Due to this mechanism, allopurinol is indicated for treating clinical manifestations associated with uric acid deposition, such as gouty arthritis, cutaneous tophi, and kidney complications involving urate crystal deposits or kidney stones. Gout, often the primary manifestation of uric acid buildup, can be aggravated by lifestyle and dietary factors. Foods high in purines, such as red meat and seafood, as well as fructose-rich fruits and vegetables, exacerbate the condition (Choi et al., 2004). Conversely, certain foods may alleviate gout symptoms; for example, cherries have been shown to be associated with lower uric acid levels and reduced gout attacks (Zhang et al., 2012). Cherries contain anthocyanins, antioxidants that help decrease inflammation. According to DrugStats, allopurinol was the 42nd most prescribed medication in the United States in 2020, with 36,600,871 prescriptions for 3,606,249 patients, moving up one rank from 2019.

A structurally similar compound, oxypurinol (Tanimoto index = 0.88), was identified as present at high concentrations in beer (Ka et al., 2006). Previous studies have shown how oxypurinol and allopurinol share xanthine oxidase as protein target (Sekine et al., 2023). Interestingly, they suggested that the mechanism of inhibition could be different in both compounds. This led us to investigate whether beer consumption, with its higher concentration of oxypurinol, might impact patients in the patients who were prescribed allopurinol. We assessed beer consumption and gout/uric acid-related symptoms to determine any correlation. Beer consumption was clearly associated with gout symptoms, particularly in terms of the number of affected joints and the duration of gout attacks, while the frequency of attacks was less significantly correlated (Figure 5A). Patients were also surveyed regarding their consumption of other foods, including red meat. Interestingly, the only significant correlation with red meat consumption was the frequency of gout attacks—the variable less associated with beer consumption (Figure 5B). This data suggests that the consumption of beer is affecting gout symptoms through a different way as the consumption of red meat, pointing to the possible effect of the presence of oxypurinol and its interaction with allopurinol.

**Figure 5.**
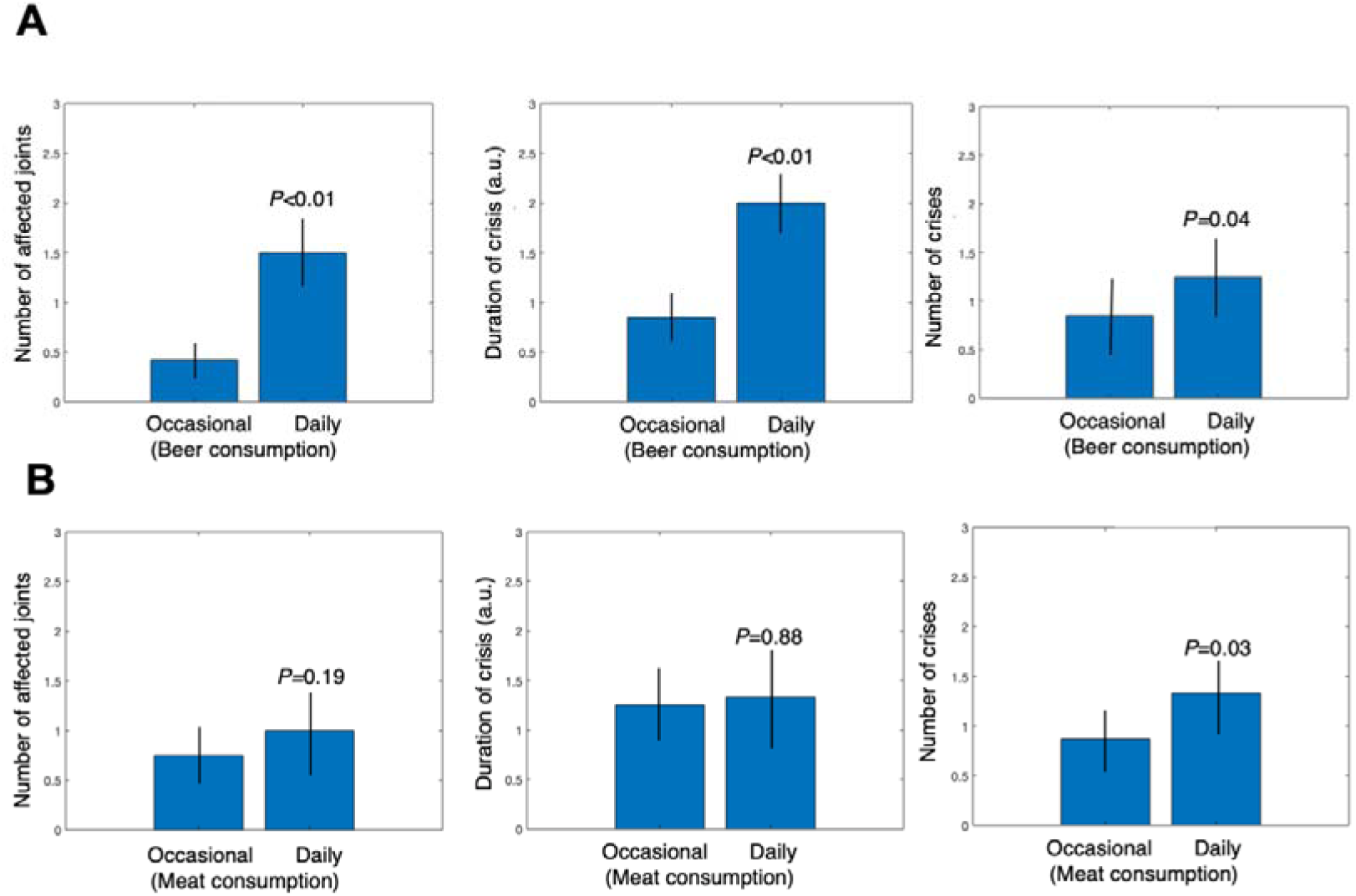
**(A)** Results from patient (allopurinol) surveys related to beer consumption and the reference of number of affected joints (left), duration of gout crises (middle) and the number of gout crises (right). **(B)** Same as in A, but for meat consumption.

## Discussion

In this paper, we developed a novel database with potential interactions between drugs and food compounds. Our hypothesis is grounded in the principle that a drug’s effect results from its interaction with one or more specific molecular targets, typically proteins, which are activated or inhibited by the drug, either through direct binding or competition with other ligands. The pharmacological response observed arises from this alteration in protein activity, as described in Goodman and Gilman’s Manual of Pharmacology and Therapeutics (Goodman and Gilman 2011), where it is noted that the effects of most drugs are mediated through their interactions with macromolecular components of the body. While our hypothesis is theoretically sound and supported by existing pharmacological frameworks (Shoichet, 2004), there are still potential limitations in our analysis that could lead to false positives within the FARFOOD database predictions.

One major limitation concerns the bioavailability of food compounds at the target site. Many ingested food compounds undergo extensive metabolism in the digestive tract and may be transformed before reaching the bloodstream or may be entirely excreted, thus never reaching the intended target cells. A solution to this limitation could involve verifying whether the predicted food compounds are indeed present in human plasma. The Human Metabolome Project (Wishart et al., 2022) is a valuable resource, containing data on over 35,000 metabolites detected in blood, and could serve as an experimental filter to validate the presence of specific compounds in circulation. By cross-referencing FARFOOD predictions with experimentally detected metabolites, we could enhance the reliability of our predictions, focusing on interactions with compounds confirmed in the bloodstream.

Another consideration is whether our selected Tanimoto index threshold (≥0.7) is restrictive enough to minimize false positives. While a higher Tanimoto index implies greater structural similarity and then a higher likelihood of interaction, it is crucial to balance sensitivity with specificity. To address this, the Tanimoto index is clearly displayed in the app interface, providing users with transparency regarding the interaction likelihood. Furthermore, the molecular docking analyses of lisinopril and bupropion to evaluate the binding affinity between the drug and its receptor in comparison with structurally similar food compounds showed a positive result. Docking studies, which are well-validated across numerous publications (Sousa et al., 2006; Trott & Olson, 2010), suggest that our computational predictions can reflect experimentally observed binding behaviors. Nonetheless, experimental validation remains necessary, and it could include *in vitro* assays where recombinant target proteins are incubated with mixtures of the drug and food compound to determine kinetic parameters and binding affinities. Since macromolecular targets are often located on cell surfaces or within specific intracellular compartments, *in vitro* testing would provide a precise measure of how food compounds influence the pharmacodynamics of drugs at the cellular level.

Moreover, our dietary habit surveys and symptom tracking for patients taking specific medications also support the existence of interactions between drugs (lisinopril, bupropion and allopurinol) and structurally similar food compounds. The survey findings align with our computational predictions and provide real-world evidence of these interactions, but it remains unclear the effect (agonist or antagonist) of the food compound with the drug. In two examples (lisinopril and bupropion), the consumption of the structurally similar food compounds correlated with positive clinical measures of the patients, while in the other example (allopurinol), the consumption of the food was associated to a negative impact in the disease.

Additionally, a promising line for future work involves developing a Polygenic Risk Score (PRS)-like approach (Kachuri et al., 2023; Kullo et al., 2022) tailored to food-drug interactions. By focusing on the target proteins listed in FARFOOD, we could identify and map the genes encoding these proteins, as genetic variations impacting these targets would likely alter the efficacy of drugs and the action of structurally similar food compounds. If we extend this analysis to food items containing multiple bioactive compounds, we could construct a PRS-like method specifically for food-related interactions. This approach would allow us to identify individuals with a high risk for a particular food, indicating a potential predisposition to altered drug responses when consuming that food. In summary, our study highlights the potential of FARFOOD as a predictive tool for identifying food-drug interactions, though further validation and refinement of these predictive methods are essential for achieving clinically relevant outcomes.

### Study Highlights

#### What is the current knowledge on the topic?

Food-Drug Interactions is a relevant and well-studied field in Pharmacology. Nevertheless, these interactions are detected in a case-by-case manner, leading to multiple undetected interactions with mild consequences in pharmacokinetics.

#### What question did this study address?

This study uses structural similarity between compounds as a method to predict potential interactions due to similar protein binding. Using a drug-wide and food-wide system, we predict multiple potential interactions between drugs and food compounds.

#### What does this study add to our knowledge?

The main result of this study is a database/application that contains the similarities (and therefore the potential interactions) between drugs and food compounds; in addition, we validate three of these predictions using *in silico* docking as well as patient surveys.

#### How might this change clinical pharmacology or translational science?

These results can lead to a deeper analysis of specific drug/food compounds in order to validate additional interactions, and then to novel nutritional recommendations for medicated patients.

## Author Contributions

A.C.R. wrote the manuscript; A.C.R. and S.M.N. designed the research; A.M.N.B. performed the research; A.M.N.B., G.C.L., D.A.B and J.M.C.G. analyzed the data;

## Data Availability

All data produced in the present study are available upon reasonable request to the authors.

http://www.github.com/acroman

## Acknowledgements

This paper was supported by the Spanish Ministry of Science through the projects TED2021-130036B-I00 and PID2020-117467RB-I00 (to SMN and ACR), and PID2021-126905NB-I00 and TED2021-130560B-I00 (to JMCG).

